# Minimal impact on the resistome of children in Botswana after azithromycin treatment for acute severe diarrhoeal disease

**DOI:** 10.1101/2023.08.03.23293510

**Authors:** Allison K. Guitor, Anna Katyukhina, Margaret Mokomane, Kwana Lechiile, David M. Goldfarb, Gerard D. Wright, Andrew G McArthur, Jeffrey M. Pernica

## Abstract

Mass distribution of azithromycin has been recommended to reduce under-five mortality rates in certain countries in sub-Saharan Africa. Additionally, antibiotic treatment of children with bacterial gastroenteritis holds promise for the prevention of mortality and the optimization of linear growth. However, mass administration and imprudent prescription of antibiotics can select for antibiotic-resistant bacteria in the gut microbiota of children. The long-term implications of this selection are unknown and worrisome.

Our previous randomized controlled trial of children hospitalized with severe acute diarrhoeal disease in Botswana evaluated the efficacy of a test-and-treat strategy. Participants randomized to the intervention group who were found to have enterotoxigenic or enteropathogenic *E. coli, Shigella,* or *Campylobacter* detectable by a rapid qualitative multiplex PCR assay at admission were treated with azithromycin and those randomized to the control group received supportive treatment (usual care). Stool samples were collected at baseline and at 60 days. In this current study, DNA from 136 stool samples was enriched and sequenced to detect changes in the resistome, otherwise known as the collection of antibiotic resistance genes.

At baseline, the gut microbiota of these children contained a diverse complement of azithromycin resistance genes that increased in prevalence in both treatment groups by 60 days. Certain 23S rRNA methyltransferases were associated with other resistance genes and mobile genetic elements, highlighting the potential for the transfer of macrolide resistance in the gut microbiome. There were other minor changes in non-azithromycin resistance genes; however, the trends were not specific to the antibiotic-treated children. In conclusion, a three-day azithromycin treatment for diarrhoea for young children in Botswana did not increase the prevalence of azithromycin-specific antibiotic resistance genes at 60 days. The gut microbiota of these children appeared primed for macrolide resistance, and repeated exposures may further select resistant bacteria.

## INTRODUCTION

In 2020, the under-five mortality rate in sub-Saharan Africa was 74 deaths per 1000 live births, totalling 2.7 million deaths, or 54% of the global value (1). Infectious diseases are associated with almost half of the global under-five deaths, with lower respiratory infections, diarrhoea, and malaria among the leading causes (2). Diarrhoea, or acute gastroenteritis, is the third-leading cause of disease burden in children under 10 years of age and is associated with significant morbidity, including cognitive maldevelopment and stunted linear growth (3–6). Viruses (i.e., rotavirus), parasites (i.e., *Cryptosporidium* spp.), and bacteria (i.e., *Shigella* spp.) are the most common aetiological agents of this disease (4). Despite decreases in childhood mortality and diarrhoeal-related deaths in the past 30 years, resulting from the implementation of oral rehydration algorithms and zinc treatment, supplemental approaches to reducing this burden are needed (1, 7, 8). Although generally reserved for the treatment of dysentery or cholera in children, recent studies have revealed that targeted treatment of bacterial gastroenteritis with the antibiotic azithromycin, perhaps through a rapid testing approach, is associated with a reduction in the duration of diarrhoea, hospitalizations, and mortality, as well as benefits in linear growth (9, 10). Additionally, the mass drug administration (MDA) of azithromycin has been recommended in certain countries with high under-five mortality rates to improve overall child survival (11).

Azithromycin, a commonly used macrolide antibiotic, targets many bacterial pathogens associated with diarrhoea and respiratory infections (12). This antibiotic also targets the bacterial-like ribosome of the apicoplast, a vestigial plastid-like organelle, rendering it effective against malarial parasites (13). There are obvious concerns with MDA and unnecessary use of an antibiotic, including unwanted impacts on the gut microbiota, selection for antibiotic-resistant organisms, and potential pressure for the mobilization and spread of antimicrobial resistance (AMR) (14–16). In 2019, over 250,000 deaths in sub-Saharan Africa were attributable to AMR, with about half occurring in children under the age of five (17). Macrolide antibiotics target the 23S rRNA of the 50S large ribosomal subunit of bacteria and interfere with mRNA translation (12). Resistance typically arises through methylation of the 23S rRNA through Erm methyltransferases, efflux, or modification of the antibiotic, or mutations to the target (18). While many studies have assessed the selection for AMR in cultured pathogens, including *Streptococcus pneumoniae* and *Escherichia coli*, few have investigated the impacts on the gut microbiota and total resistome (i.e., collection of all antibiotic resistance genes (ARGs)) (14, 19–23). It is imperative to weigh the benefits of MDA of azithromycin and its indication to treat bacterial gastroenteritis against potential impacts on selection for AMR in the long term.

We completed a randomized controlled trial (RCT) of children with severe acute diarrhoeal disease in southern Botswana, comparing a rapid test-and-treat strategy (resulting in azithromycin treatment for those with treatable enteric bacterial pathogens) against supportive care (usual treatment) (10, 24). We hypothesized that azithromycin exposure would select for an increase in the prevalence and abundance of macrolide ARGs in the gut microbiome of these children compared to those that received the usual care. Metagenomic DNA from stool samples before and 60 days after treatment was assayed with a targeted capture method to selectively sequence ARGs. The azithromycin-specific and total gut resistome of these children were assessed to identify potential consequences of antibiotic exposure.

## METHODS

### Study design and participants for this sub-study

A full description of the multicentre, randomised, controlled trial has previously been published (10). Children hospitalized because of severe acute diarrhoea were randomised to either the Rapid Test-and-Treat (RTT) or Usual Care (UC) only group. All study participants received usual care, consisting of fluid rehydration and zinc treatment for children with acute diarrhoeal disease as per World Health Organization standards (25); those in the RTT group also received this intervention. Each group was further randomized to either receive a probiotic supplementation of *Lactobacillus reuteri* DSM 17938 (1×10^8^ colony forming units by mouth) once daily or a placebo for 60 days (10). In the RTT group, rectal swabs were assayed using a multiplex PCR-based qualitative panels, and those with enteric specimens positive for *Shigella*, enterotoxigenic *Escherichia coli* (ETEC), enteropathogenic *E. coli* (EPEC), and/or *Campylobacter*, were treated orally with azithromycin (10 mg/kg once daily for three days). Those positive for *Cryptosporidium* were treated with nitazoxanide. A subset of 34 children from the RTT group that received azithromycin treatment and 34 children from the UC group were selected for this study. Bulk stool samples were collected at baseline (before treatment) and 60 days after enrolment and kept at −80°C until further processing.

### Targeted sequencing of antibiotic resistance genes

DNA was extracted from 0.1 – 0.2 g of stool as previously described (26, 27). Full details on dsDNA library preparation and enrichment for ARGs are described in Supplementary Methods. Enrichment for ARGs was performed as previously described using a probe set to target over 2,000 ARGs (28, 29). Enriched libraries were sequenced on an Illumina MiSeq with 2 x 300 bp sequencing chemistry to a targeted depth of 250,000 clusters per library. Ten negative controls of a buffer-only extraction blank were processed alongside the stool samples and five were sequenced after enrichment.

### Analysis of captured antibiotic resistance genes

A full description of the analysis is described in the Supplementary Methods. After subsampling reads, we used two analysis modules from the Resistance Gene Identifier (RGI): a read-mapping approach (RGI*bwt) and a *de novo* assembly and ARG prediction method (RGI*main). The former method estimates the number of reads that map to ARG sequences in the Comprehensive Antibiotic Resistance Database (CARD) v 3.2.1 (30, 31). This method has difficulty resolving the specific variants of members within large AMR gene families (AGFs) but can infer the abundance of ARGs. AGFs are a higher classification of ARGs; for example, *ermF* (ARO: 3000498) and *ermG* (ARO: 3000522) are both members of the “Erm 23S ribosomal RNA methyltransferase” AGF in CARD. The latter method (RGI*main) can better resolve variants but cannot inform on the abundance of ARGs and relies on the assembly of contiguous sequences of DNA (contigs) of sufficient length to predict ARGs through comparison to CARD sequences.

We compared results between the two treatment groups at the ARG and the AGF levels. The prevalence of certain ARGs was determined from the RGI*main results and their abundance was estimated from the RGI*bwt results. For the 23S rRNA methyltransferases detected through RGI*bwt, the prevalence of predicted bacterial hosts from the RGI*kmer_query for prediction of pathogen of origin analysis module was compared across cohorts and timepoints. Finally, the surrounding genetic context of these methyltransferases, along with the macrolide phosphotransferases, was used to infer potential bacterial hosts and co-localized ARGs.

### Patient and public involvement

Patients and the public were not involved in the design, or conduct, or reporting, or dissemination plans of this research.

## RESULTS

### Patient characteristics

Our study focused on a sample of 68 children from the original RCT (10) (Table 1). Approximately 50% of children in each arm of the study received *L. reuteri* probiotic for 60 days. Most children were positive for either EPEC or ETEC, and a small percentage (8/34) were positive for *Campylobacter* spp. Three children tested positive for *Cryptosporidium*, in addition to a bacterial pathogen, and received nitazoxanide in addition to azithromycin. Many children in both arms of the study received antibiotics before enrolment, including cefotaxime, amoxicillin, co-trimoxazole, and ampicillin (Additional File 1). One child in the SC group received erythromycin before enrolment.

**Table 1.** Participant characteristics of children included in the resistome study.

| Factor | Usual Care<br>(n = 34) | Rapid Test-and-Treat<br>(n = 34) |
| --- | --- | --- |
| Male, N (%) | 17 (50.0%) | 20 (58.8%) |
| Age (yrs) at baseline<br>(average +/- SD) | 0.988 +/- 0.736 | 0.995 +/- 0.505 |
| Probiotic supplementation, N (%) | 16 (47.0%) | 16 (47.0%) |
| Prior antibiotic exposure, N (%) | 14 (41.2%) | 7 (20.6%) |
| *Campylobacter* positive, N (%) | Not tested | 8 (23.5%) |
| *Shigella* positive, N (%) | Not tested | 4 (11.8 %) |
| *Cryptosporidium* positive, N (%) | Not tested | 3 (8.8%) |
| Enterotoxigenic *E. coli* positive, N (%) | Not tested | 11 (32.4%) |
| Enteropathogenic *E. coli* positive, N (%) | Not tested | 25 (73.5%) |
| Other pathogen positive, N (%) | Not tested | 22 (64.7%) |
| Azithromycin exposure, N(%) | 0 (0%) | 34 (100%) |

### Capturing the resistome of children treated with azithromycin

Using a method to selectively target and sequence over 2,000 ARGs from metagenomic DNA extracted from stool, we detected, on average, between 76 to 81 ARGs per child through read mapping to CARD, and slightly fewer through the more stringent method of *de novo* assembly and RGI (66 to 75 ARGs on average per child) (Figure S1BD). Overall, we found no appreciable differences in the number of ARGs or AGFs between the two cohorts of children. Through the *de novo* assembly and RGI*main analysis, the number of ARGs per child increased on average by 5.2 genes for the UC cohort and 6.9 for the RTT cohort at 60 days (Figure S1D). We found the increase in ARGs to be significant only in the azithromycin-treated group (Figure S1D). In general, the number of AGFs also increased from baseline to 60 days in both cohorts. We reported an average increase of 3.8 (RGI*bwt) or 3.3 (RGI*main) AGFs for the RTT children and 2.4 (RGI*bwt) or 1.8 (RGI*main) AGFs for the UC children. This increase was significant only in azithromycin-treated children (Figure S1CE). There were many ARGs that were unique to one group; however, these were typically rare ARGs in the entire group and only found in a few individuals (Figures S2AB, S3AB; Additional Files 2, 3). Despite the individual differences in AGFs identified in azithromycin-treated children by 60 days, there were very few families that were unique to one group, and these were often only identified in one or two children (Figures S2CD, S3CD; Additional Files 2, 3).

### Increase in the prevalence of macrolide resistance genes in both cohorts

At baseline, the macrolide resistome appeared similar across azithromycin-treated infants and those who received usual care (Figures 1A, S4). There were distinct changes in the prevalence and abundance of 23S ribosomal RNA methyltransferases that confer resistance to macrolide, lincosamide, and streptogramin antibiotics. In both cohorts, the prevalence of *ermQ*, *ermF*, *ermT*, *ermX*, and *ermG* increased at 60 days by 13% to 55% (RGI presence/absence results) (Figure 1). Many children maintained these genes from baseline to 60 days, while in some children, they appeared during the study period (Figure 1B). We found no significant differences in the prevalence of these genes between cohorts at 60 days (Figure 1A). Before antibiotic treatment, the children in the RTT arm had more reads mapping to *ermF* (ARO: 3000498) than those in the UC arm (Figure S5A). While both cohorts experienced an increase in reads mapping to *ermF* (average of 2259 for RTT and 1376 for UC), this gene remained more abundant at 60 days in the azithromycin-treated children (average of 4252 reads in RTT vs 2363 in UC) (Figure S5). At 60 days, on average, more reads were mapped to *ermT* (ARO: 3000595) in the UC group (3348 for UC vs 2128 for RTT), which had increased from baseline (average reads of 1792 for UC vs 1757 for RTT) (Figure S5). Whereas *ermX* (ARO: 3000596) read counts increased in the azithromycin-treated children by 60 days (average of 5409 reads and an increase of 3554), this gene was slightly decreased (average of 79 reads) in UC children at 60 days (Figure S5).

**Figure 1:**
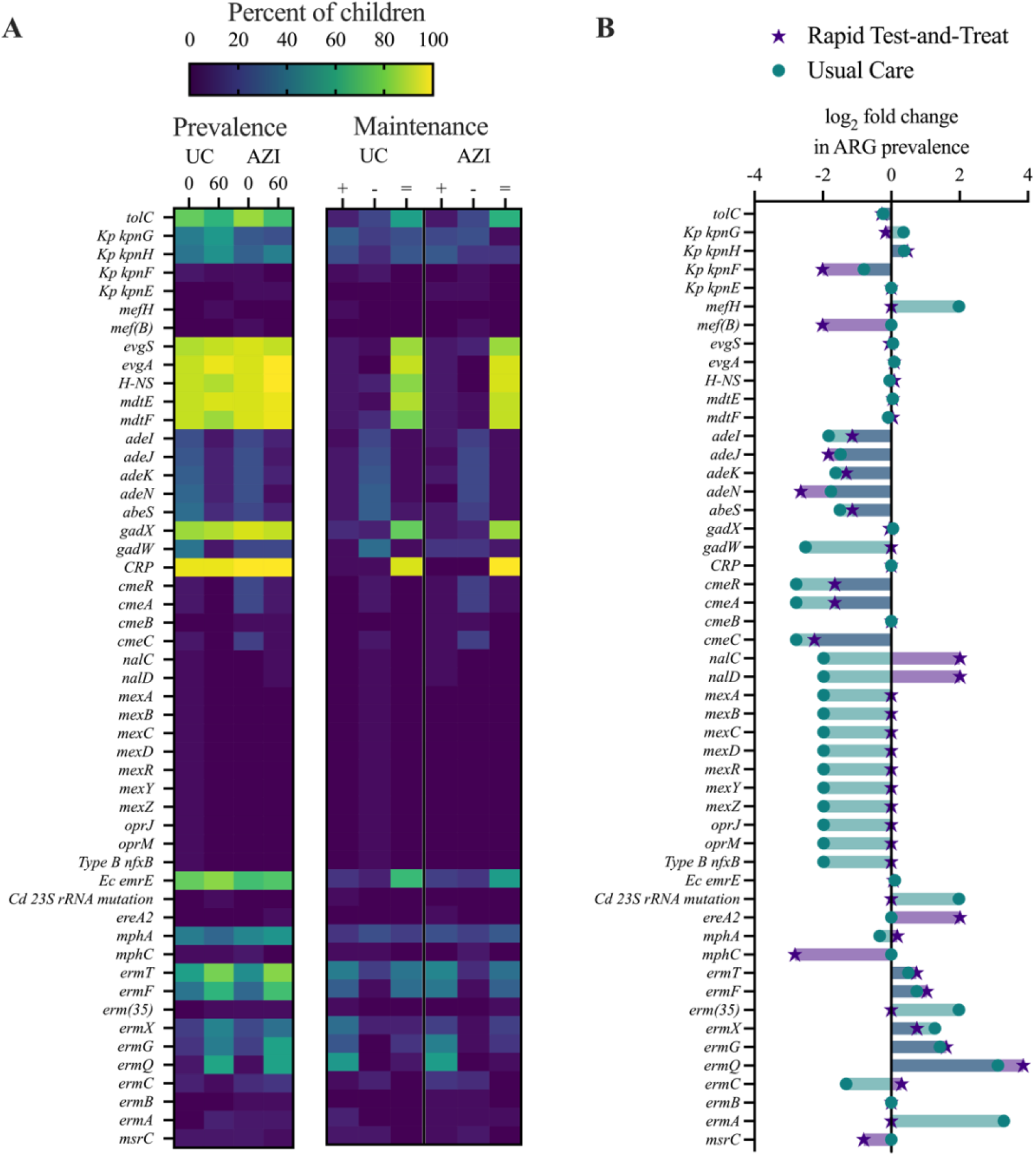
Prevalence of macrolide resistance genes in infants and changes at 60 days. A) Heatmap showing the prevalence and maintenance of macrolide ARGs in each group of treated children. In the left panel, the values correspond to the percentage of children in each group at each time point for which a given ARG was detected through *de novo* assembly and RGI*main analysis. The right panel shows the percentage of children in which a gene appeared (+), disappeared (-) or was maintained (=) by 60 days in each treatment group. B) The log_2_ fold change in macrolide ARG prevalence by 60 days in both groups. UC = Usual Care, AZI = Rapid Test-and-Treat, Kp = *Klebsiella pneumoniae*, Ec = *Escherichia coli*, Cd = *Clostridioides difficile*.

In addition to the ribosomal methyltransferases, there were few changes in other genes implicated in macrolide resistance (Figures S4, S5). At baseline, the macrolide phosphotransferase *mphA* (ARO: 3000316) was more prevalent in the RTT arm (46%) than the UC arm (38%) and remained so at 60 days (50% for RTT vs 32% for UC) (Figure 1A). Finally, certain efflux systems (e.g., *cme* genes found in *Campylobacter* spp. and *ade* genes originating in *Acinetobacter* spp.) were reduced in prevalence in both groups by 60 days (Figure 1, S4, S5).

### Potential bacterial hosts of macrolide resistance genes

Given the rising frequency of 23S rRNA methyltransferases in both cohorts, we next predicted the bacterial hosts of these ARGs, which were similar between the azithromycin-treated children and those that received usual care (Figure 2). *Bacteroides* spp. were common for both *ermF* and *ermG*, while *ermX* likely originated in *Bifidobacterium* spp. or *Corynebacterium* spp. (Additional File 4). Interestingly, in both groups of children at 60 days, *ermG* was predicted in the additional host *Klebsiella* spp.. A more diverse range of bacteria was detected for *ermT,* including *Enterococcus faecium, Escherichia* spp*., Klebsiella* spp., *Staphylococcus* spp. and *Streptococcus suis*. Additional potential hosts identified through the BLAST analysis included *Clostridiodes* spp. for *ermG,* and *Intestinibacter* spp., *Peptacetobacter* spp., or *Clostridium perfringens* for *ermQ* (Additional File 4). We further annotated the genetic context of these genes using Prokka and identified *ermTFX* alongside insertion sequences (IS) in some children (Figure 3; Additional File 4).

**Figure 2:**
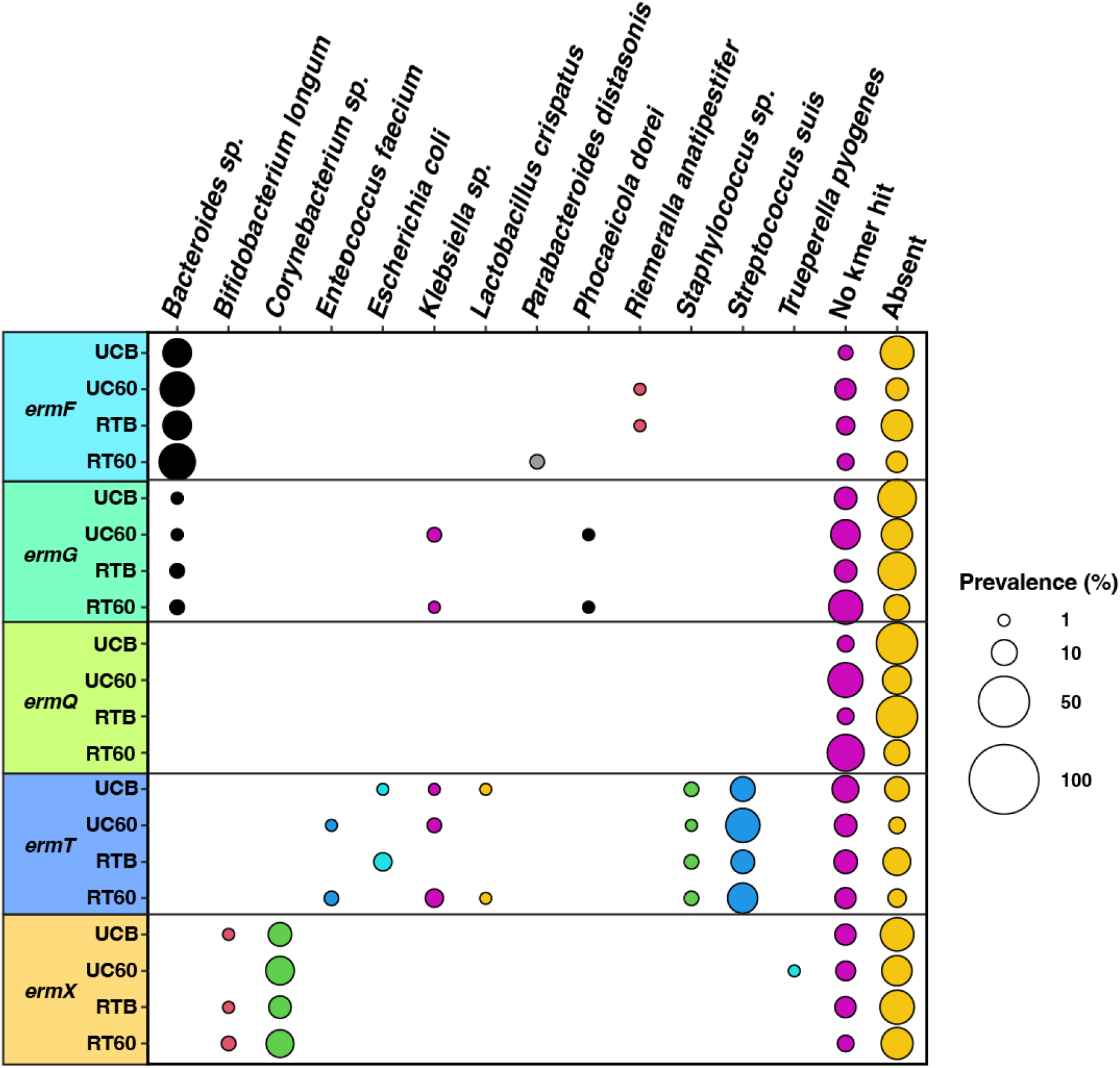
Predicted bacterial hosts of selected macrolide resistance genes in infants. Bacterial hosts were predicted through RGI*kmer_query from the *de novo* assembly and RGI*main results. Each bubble represents the percentage of children in each cohort at a given time point for which a specific host was predicted for a given ARG. “No kmer hit” signifies that the ARG did not have any significant kmer matches with the current database. “Absent” signifies the percentage of children in which the ARG gene not identified through the RGI*main analysis. Only the *erm* ARGs with distinct changes in prevalence or abundance are shown. UCB = Usual Care Baseline, UC60 = Usual Care 60 days, RTB = Rapid Test-and-Treat Baseline, RT60 = Rapid Test-and-Treat 60 days.

**Figure 3:**
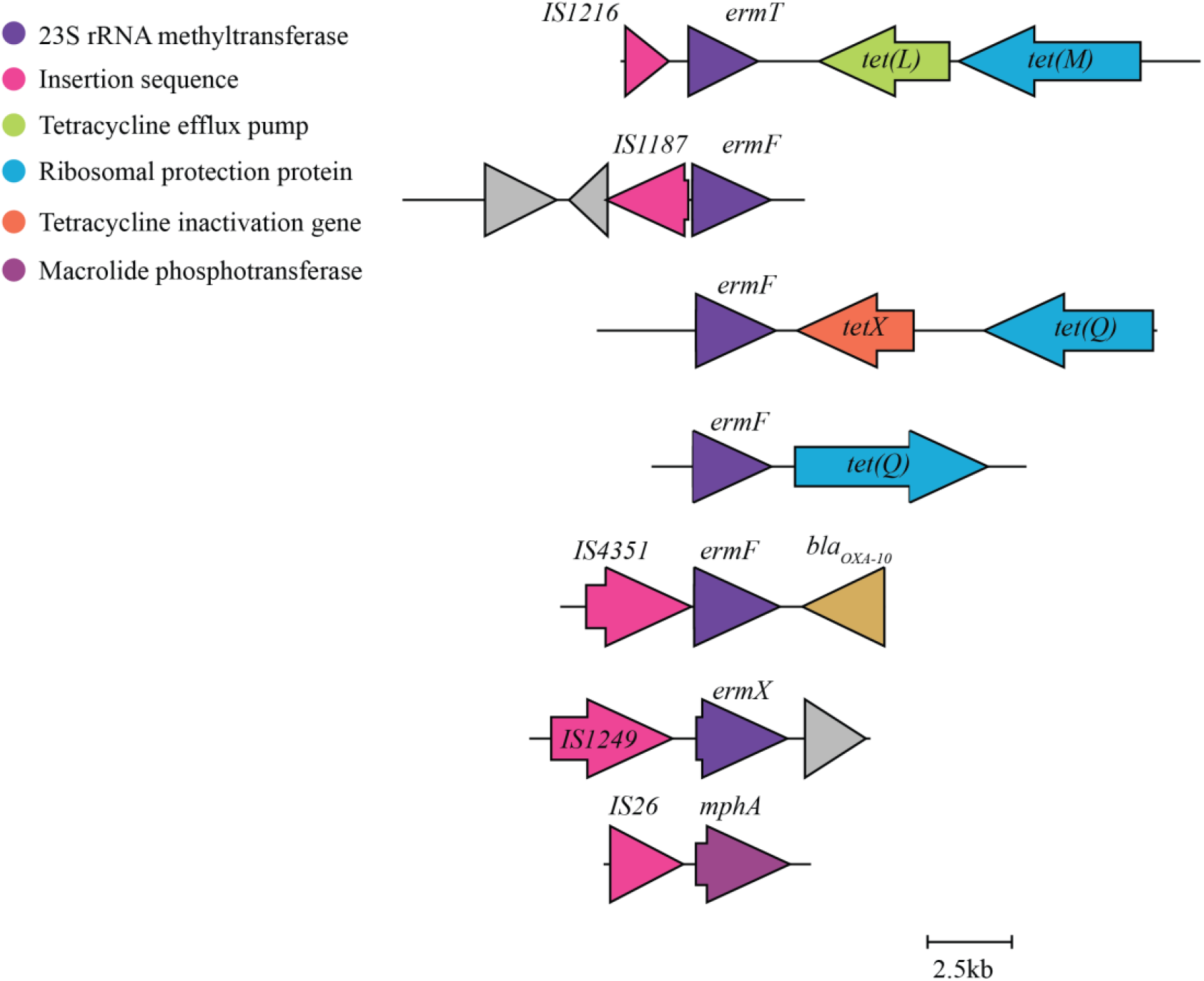
Representative genetic context of selected macrolide resistance genes in infants. Contigs obtained through *de novo* assembly and predicted to contain various macrolide resistance genes were annotated using Prokka. Any ARGs present within the same contig were identified through RGI*main. While a variety of genetic contexts were observed in both the Usual Care and Rapid-Test-and-Treat cohorts, a representative contig that was found in both groups is shown here as an example. Other contexts are described in Additional File 4.

### Changes in non-macrolide resistance genes

Next, we sought to compare differences in ARGs not associated with macrolide resistance (Figures 4, S6). In general, the gut microbiome of children in both cohorts acquired trimethoprim-resistant dihydrofolate reductase (*dfr*) genes by the 60-day follow-up and the abundance of *dfrF* increased at 60 days in both groups (Figures 4, S7). After 60 days, more azithromycin-treated children acquired the beta-lactamase *bla_cfxA3_,* and in both cohorts there was a slight increase in the number of reads mapping to members of the *bla*_cfxA_ family in general (Figures 4, S7). Although we did not find any significant differences in the average number of reads mapping to members of the following AGFs, they appear to change in prevalence over time (Figures 4, S6): vancomycin resistance genes belonging to the *vanG* and *vanC* clusters, and the *cepA* beta-lactamases appear in the gut microbiome of more children in both groups by 60 days, whereas the prevalence of genes belonging to the *bla*_OXA_ gene family was reduced in the gut microbiome of children in both groups by 60 days (Figure 4).

**Figure 4:**
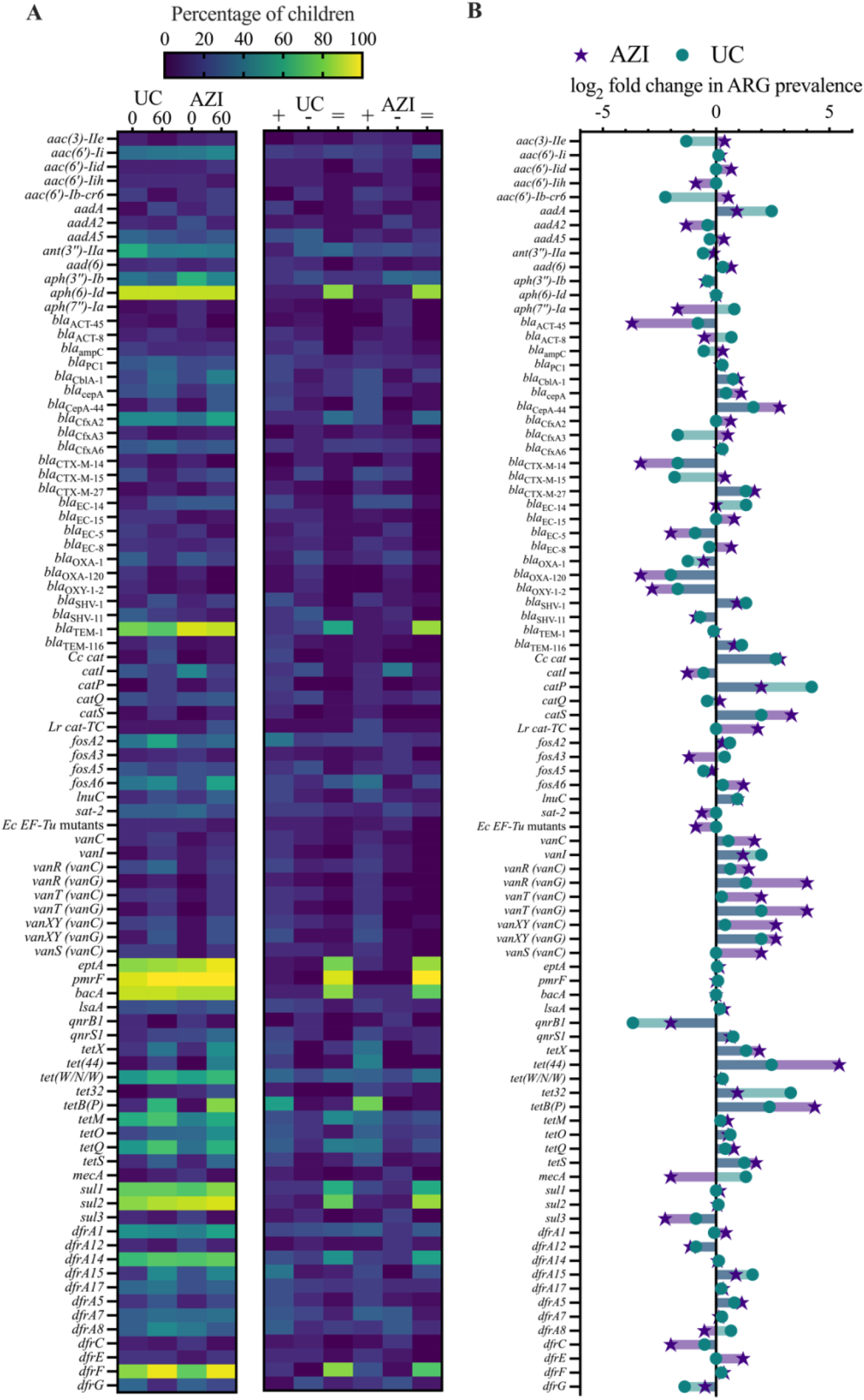
Prevalence of ARGs present in at least 10% of infants in at least one time point and changes at 60 days. A) Heatmap showing the prevalence and maintenance of ARGs in each group of treated children. In the left panel, the values correspond to the percentage of children in each group at each time point for which a given ARG was detected through *de novo* assembly and RGI*main analysis. The right panel shows the percentage of children in which a gene appeared (+), disappeared (-) or was maintained (=) by 60 days in each treatment group. B) The log_2_ fold change in macrolide ARG prevalence by 60 days in both groups. Macrolide ARGs were omitted because they are included in Figure 1. UC = Usual Care, AZI = Rapid Test-and-Treat, Ec = *Escherichia coli*, Lr = *Limosilactobacillus reuteri,* Cc *= Campylobacter coli*.

From baseline to 60 days, there was an increase in the prevalence and abundance of tetracycline-resistant ribosomal protection proteins (*tetO*, *tetQ*, *tetS*, *tetB*(P), *tet32*; ARO: 3000190, 3000191, 300192, 3000195, 3000196) in both groups (Figures 4, S6, S7). The gene, *tetX* (ARO:3000205), encoding a tetracycline inactivating monooxygenase, also increased in prevalence and abundance in both groups by 60 days (from 19% to 38% in the UC group and 14% to 44% in the RTT children) (Figures 4, S6, S7). When we assessed the genetic context of the Erm methyltransferases, we found that *ermT* was commonly associated with *tetM* and tetracycline efflux pumps, including *tet*(*45*) *and tet(L)*. We also identified *ermF* on assembled DNA fragments containing *tetQ* and *tetX* (Figure 3).

## DISCUSSION

Azithromycin holds the promise to prevent child mortality and improve linear growth in children living in low- and middle-income country settings, especially in those with acute diarrhoeal disease; however, the question of whether these benefits would be obviated by the emergence of resistance has repeatedly been raised (9, 32). Our study is, to our knowledge, the first to comprehensively investigate the effect of azithromycin on the gut resistome of children with diarrhoea in sub-Saharan Africa. Although the children that received azithromycin experienced a greater increase in the total number of AGFs by 60 days, this was not associated with a difference in macrolide resistance genes when compared to the children that received the usual treatment for diarrhoea. In both groups, certain macrolide- and non-macrolide ARGs persisted or increased in prevalence regardless of antibiotic exposure. Finally, certain macrolide resistance determinants were linked to insertion sequences and other ARGs highlighting the potential for dissemination and spread of AMR within the gut microbiome of these children. In this survey of children under five years of age, we revealed the diversity of the gut resistome and showed that a three-day exposure to azithromycin did not provide additional pressure to retain macrolide resistance genes two months later.

We did not find a significant increase in the total number of ARGs from baseline to 60 days for either group of infants. There was a general increase in the number of AGFs in both groups, although this was only significant in the azithromycin-treated children and may reflect small inter-individual differences. Given that these children presented to the hospital with acute gastroenteritis, their gut microbiota may have been dominated by the infection-causing organism at this time (33, 34). Most of the tested children in our sub-study were infected by a pathogenic *E. coli*, either EPEC or ETEC, and many likely experienced co-infections. Many children in both groups also received other antibiotic treatments as part of usual care. It is possible that by 60 days the gut microbiota would have recovered from the infection or was in the process of being restored to a more diverse ecosystem where members of the genera *Bacteroides* and *Prevotella* were more abundant (33, 35). Although we did not measure changes in the microbial diversity of these children from baseline to 60 days, the microbiota likely transformed dramatically throughout recovery (35, 36). The increase in AGFs we observed may reflect an increase in commensal and other beneficial bacteria of the gut microbiota that encode a variety of intrinsic ARGs distinct from those of diarrhoeal-causing pathogens (37).

Focusing on macrolide resistance genes, we identified a diversity of Erm 23S rRNA methyltransferases in both groups of children. At baseline, these genes were similar in prevalence in both groups of children and increased by 60 days, irrespective of the treatment received. Data on the prevalence of macrolide resistance in children in Botswana is limited; however, a recent study of wastewater found high levels of resistant bacteria in the environment (38). Although the abundance of certain *erm* genes changed more dramatically in one treatment group compared to the other, there is large individual variability within each group that may have skewed the results. By analyzing the genetic surroundings of these *erm* genes, we inferred they were likely encoded by bacteria often found in the gut microbiota and involved in the general restoration of the microbiome after a diarrhoeal infection; therefore, changes in *erm* genes in either treatment group are not likely related to an increase in diarrhoeal-causing bacteria in the gut microbiome (35, 36).

A macrolide phosphotransferase, encoded by the gene *mphA*, was also prevalent in these children upon hospitalization. This gene slightly decreased in prevalence in the UC group of children and increased in the azithromycin-treated children; however, it was also more prevalent in this latter group at baseline. Therefore, it is difficult to associate the increase in prevalence with azithromycin treatment alone. What is concerning, however, is the association of this macrolide phosphotransferase with IS elements. Plasmids containing *mphA* have been identified in ETEC isolates and are common in bacteria circulating in wastewater and river environments in Botswana (38, 39).

Finally, many of the *erm* genes identified in this group of children were commonly associated with insertion sequences, mobile elements, and tetracycline resistance genes. Although both tetracycline ARGs and *erm* genes are often found in commensal gut bacteria, their ability to transfer within genera to potential pathogenic strains and risk of co-selection is worrisome (40). Tetracyclines and macrolides are prescribed less often than other antibiotics such as cefotaxime and metronidazole in Botswana (41). Nevertheless, repeated exposure to either antibiotic might put these young children at future risk for the transfer of ARGs within their gut microbiota and the persistence of resistant strains.

Our study focused on a three-day treatment of diarrhoea with azithromycin. In contrast, the MORDOR trials studying the MDA of azithromycin to reduce mortality in children assessed the impact of repeated exposures on resistance. They found longer-term impacts on the gut resistome and microbiome, including decreased microbial diversity, increased macrolide gene expression, and macrolide resistance in *S. pneumoniae* after 2 years of biannual azithromycin administration (20–23). Finally, after 4 years of biannual exposure, they observed the selection of non-macrolide resistance genes towards aminoglycosides, beta-lactams, trimethoprim, and metronidazole (21). Although these studies used the more comprehensive method of metagenomic shotgun sequencing they relied on pooled samples for their analysis, which could be biased by a few individuals and cannot resolve changes at the individual-level (21, 22). The disparities between our results and those of these trials on biannual distribution may relate to the resilience of the gut microbiome to short-term courses of antibiotics compared to repeated exposures and other differences in trial design (42).

The short-term use of azithromycin for the treatment of acute diarrhoea in young children has also been assessed in a multi-country RCT where participants were randomized to receive azithromycin or placebo (19). Similar to our findings, the ABCD study group did not identify differences in antibiotic susceptibilities in bacterial isolates from participants that received azithromycin as compared to those received placebo(19). The main trial was stopped for futility since mortality was much lower than expected in both treatment groups; however, a retrospective sub-group analysis focusing on only those likely to have had bacterial enteritis found that azithromycin receipt was associated with improvements in linear growth and less persistent diarrhoea, hospitalization, and mortality (9). Their results highlight the potential significant benefits of targeted antibiotic therapy when the aetiological agent of acute gastroenteritis is known to be bacterial. Our current analysis of the impact on the gut resistome is suggestive that these important gains may be had without a significant increase in AMR, at least in the short term.

It remains imperative to understand baseline levels of antibiotic resistance, rapidly detect pathogens in infections, and to perform longitudinal sampling to fully characterize the changes that occur in the gut resistome post-antibiotic exposure. Shotgun metagenomics is still a relatively expensive approach and may miss rare members of the resistome. A targeted sequencing method, such as the one used in our study, can capture over 2,000 ARGs with minimal sequencing depth and cost and provides sensitive and high-quality resolution of the entire resistome. One limitation is the inability to capture resistance due to mutations in target genes (i.e., 23S rRNA mutations). These mutations may be selected rapidly after azithromycin treatment and would be missed in our analysis. Bacteria encode variable copies of the 23S rRNA locus *rrn*; therefore, the extent of resistance due to mutations depends on the number of modified copies and may not be as relevant as other resistance mechanisms (18). Another limitation is that we only assessed stool samples at baseline and 60 days and, therefore, may have missed changes in the gut resistome of these children immediately after azithromycin treatment and in the longer term.

Although a short three-day course of azithromycin for diarrhoea did not have an appreciable selective effect on the resistome of children in our study, repeated exposures, such as MDA, risk further selection in an environment primed with resistance. We were surprised to identify such a diverse set of macrolide resistance genes in children at a young age and that these generally increased in prevalence in both cohorts regardless of the treatment they received. Bystander exposure to antibiotics, similarities in the recovery of the gut microbiome after diarrhoea in both groups, and other unknown factors may have contributed to this result (43). While the macrolide ARGs identified in this study were predominantly associated with gut commensal organisms, horizontal gene transfer to potential pathogens is still a concern. Despite azithromycin being a relatively cheap and effective intervention to reduce childhood mortality and treat bacterial gastroenteritis, we believe the unknowns associated with selection for antibiotic resistance, the linkage of these resistance genes with mobile elements, and the early warnings from other trials on resistance, warrant investigating alternative interventions and rapid diagnostics. The incidence of diarrhoea and other infectious diseases may be reduced through vaccination, access to safe drinking water, nutrient supplementation, improved sanitation, and access to quality health care (4, 8, 44–47). These interventions should be further explored to prevent the MDA of antibiotics as the longer-term impacts of antibiotic treatment on the resistome of children have yet to be fully explored.

## Data Availability

Sequence data that support the finding of this study have been deposited in NCBI's Sequence Read Archive with BioProject accession code: PRJNA980291 and will be available upon publication. Code used to analyze the data is available at: https://github.com/AllisonGuitor/AMR-metatools.

## ABBREVIATIONS

23S rRNA: 23S ribosomal ribonucleic acid
AGF: Antimicrobial resistance gene family
AMR: Antimicrobial resistance
ARG: antibiotic resistance gene
ARO: Antibiotic Resistance Ontology
CARD: Comprehensive Antibiotic Resistance Database
DNA: deoxyribonucleic acid
EPEC: Enteropathogenic Escherichia coli
ETEC: Enterotoxigenic Escherichia coli
MDA: mass drug administration
PCR: polymerase chain reaction
qPCR: quantitative PCR
RGI: Resistance Gene Identifier
RTT: Rapid Test-and-Treat
UC: Usual Care

## CONTRIBUTIONS

The randomized controlled trial from which the samples were selected was led by MM, DMG, and JMP; MM and KL oversaw stool testing and storage. AGM, JMP, DMG, GDW, and AKG conceived the study and designed the experiments. AKG and AK performed the DNA extraction, library preparation, and targeted enrichments. AKG analyzed the data, generated tables and figures, and wrote the manuscript with primary input from AGM, GDW, and JMP. All authors read and approved the final manuscript.

### ACKNOWLEDGMENTS

We acknowledge Michelle Shah for help with DNA extractions and the McMaster Genomics Facility for next-generation sequencing. We thank Ms. Banno Moorad for their contributions towards data collection and Dr. David Speicher (Redeemer University, Canada) for assistance in the early stages of planning this work.

## COMPETING INTERESTS

JMP’s institution has received grant funding from MedImmune for a study outside the submitted work.

**Patient and public involvement** Patients and/or the public were not involved in the design, or conduct, or reporting, or dissemination plans of this research.

**Patient consent for publication.** Not applicable.

## Ethics approval

This study involves human participants and was approved by the Hamilton Integrated Research Ethics Board, the Botswana Ministry of Health and Wellness Health Research Development Committee Institutional Research Board, and the Princess Marina Hospital Research Ethics Committee. Participants gave informed consent to participate in the study before taking part.

## AVAILABILITY OF DATA AND MATERIALS

Sequence data that support the finding of this study have been deposited in NCBI’s Sequence Read Archive with BioProject accession code: PRJNA980291. Code used to analyze the data is available at: https://github.com/AllisonGuitor/AMR-metatools.

## REFERENCES

1. UNICEF. Levels & Trends in Child Mortality - Report 2021 [online]. 2021. Available from: https://data.unicef.org/resources/levels-and-trends-in-child-mortality-2021/ (accessed November 1, 2022).

2. Perin J, Mulick A, Yeung D, et al. Global, regional, and national causes of under-5 mortality in 2000–19: an updated systematic analysis with implications for the Sustainable Development Goals. The Lancet Child & Adolescent Health. 2022;6(2):106–15; doi:10.1016/s2352-4642(21)00311-4.

3. Vos T, Lim SS, Abbafati C, et al. Global burden of 369 diseases and injuries in 204 countries and territories, 1990 - 2019: a systematic analysis for the Global Burden of Disease Study 2019. The Lancet. 2020;396(10258):1204–22; doi:10.1016/S0140-6736(20)30925-9.

4. Troeger C, Forouzanfar M, Rao PC, et al. Estimates of global, regional, and national morbidity, mortality, and aetiologies of diarrhoeal diseases: a systematic analysis for the Global Burden of Disease Study 2015. The Lancet Infectious Diseases. 2017;17(9):909–48; doi:10.1016/S1473-3099(17)30276-1.

5. Nasrin D, Blackwelder WC, Sommerfelt H, et al. Pathogens Associated With Linear Growth Faltering in Children With Diarrhea and Impact of Antibiotic Treatment: The Global Enteric Multicenter Study. J Infect Dis. 2021;224(12 Suppl 2):S848-S55; doi:10.1093/infdis/jiab434.

6. Rogawski ET, Liu J, Platts-Mills JA, et al. Use of quantitative molecular diagnostic methods to investigate the effect of enteropathogen infections on linear growth in children in low-resource settings: longitudinal analysis of results from the MAL-ED cohort study. Lancet Glob Health. 2018;6(12):e1319–e28; doi:10.1016/S2214-109X(18)30351-6.

7. Mokomane M, Kasvosve I, de Melo E, et al. The global problem of childhood diarrhoeal diseases: emerging strategies in prevention and management. Ther Adv Infect Dis. 2018;5(1):29–43; doi:10.1177/2049936117744429.

8. Malande OO, Munube D, Afaayo RN, et al. Barriers to effective uptake and provision of immunization in a rural district in Uganda. PLoS One. 2019;14(2):e0212270; doi:10.1371/journal.pone.0212270.

9. Pavlinac PB, Platts-Mills J, Liu J, et al. Azithromycin for bacterial watery diarrhea: A reanalysis of the AntiBiotics for Children with severe Diarrhea (ABCD) trial incorporating molecular diagnostics. J Infect Dis. 2023; doi:10.1093/infdis/jiad252.

10. Pernica JM, Arscott-Mills T, Steenhoff AP, et al. Optimising the management of childhood acute diarrhoeal disease using a rapid test-and-treat strategy and/or *Lactobacillus reuteri* DSM 17938: a multicentre, randomised, controlled, factorial trial in Botswana. BMJ Glob Health. 2022;7(4); doi:10.1136/bmjgh-2021-007826.

11. WHO. WHO guideline on mass drug administration of azithromycin to children under five years of age to promote child survival [online]. 2020. Available from: https://apps.who.int/iris/handle/10665/333942 (accessed November 1, 2022).

12. Parnham MJ, Erakovic Haber V, Giamarellos-Bourboulis EJ, et al. Azithromycin: mechanisms of action and their relevance for clinical applications. Pharmacol Ther. 2014;143(2):225–45; doi:10.1016/j.pharmthera.2014.03.003.

13. Burns AL, Sleebs BE, Gancheva M, et al. Targeting malaria parasites with novel derivatives of azithromycin. Front Cell Infect Microbiol. 2022;12:1063407; doi:10.3389/fcimb.2022.1063407.

14. O’Brien KS, Emerson P, Hooper PJ, et al. Antimicrobial resistance following mass azithromycin distribution for trachoma: a systematic review. The Lancet Infectious Diseases. 2019;19(1):e14–e25; doi:10.1016/s1473-3099(18)30444-4.

15. Keenan JD, Arzika AM, Lietman TM. Azithromycin and Childhood Mortality in Africa. New England Journal of Medicine. 2018;379(14):1382–4; doi:10.1056/NEJMc1808346.

16. Hooda Y, Tanmoy AM, Sajib MSI, et al. Mass azithromycin administration: considerations in an increasingly resistant world. BMJ Glob Health. 2020;5(6); doi:10.1136/bmjgh-2020-002446.

17. Murray CJL, Ikuta KS, Sharara F, et al. Global burden of bacterial antimicrobial resistance in 2019: a systematic analysis. The Lancet. 2022;399(10325):629–55; doi:10.1016/s0140-6736(21)02724-0.

18. Gomes C, Martinez-Puchol S, Palma N, et al. Macrolide resistance mechanisms in Enterobacteriaceae: Focus on azithromycin. Crit Rev Microbiol. 2017;43(1):1–30; doi:10.3109/1040841X.2015.1136261.

19. Antibiotics for Children With Diarrhea Study G, Ahmed T, Chisti MJ, et al. Effect of 3 Days of Oral Azithromycin on Young Children With Acute Diarrhea in Low-Resource Settings: A Randomized Clinical Trial. JAMA Netw Open. 2021;4(12):e2136726; doi:10.1001/jamanetworkopen.2021.36726.

20. Arzika AM, Maliki R, Abdou A, et al. Gut Resistome of Preschool Children After Prolonged Mass Azithromycin Distribution: A Cluster-randomized Trial. Clin Infect Dis. 2021;73(7):1292–5; doi:10.1093/cid/ciab485.

21. Doan T, Worden L, Hinterwirth A, et al. Macrolide and Nonmacrolide Resistance with Mass Azithromycin Distribution. N Engl J Med. 2020;383(20):1941–50; doi:10.1056/NEJMoa2002606.

22. Doan T, Arzika AM, Hinterwirth A, et al. Macrolide Resistance in MORDOR I — A Cluster-Randomized Trial in Niger. New England Journal of Medicine. 2019;380(23):2271–3; doi:10.1056/NEJMc1901535.

23. Doan T, Hinterwirth A, Worden L, et al. Gut microbiome alteration in MORDOR I: a community-randomized trial of mass azithromycin distribution. Nat Med. 2019;25(9):1370–6; doi:10.1038/s41591-019-0533-0.

24. Pernica JM, Steenhoff AP, Mokomane M, et al. Rapid enteric testing to permit targeted antimicrobial therapy, with and without *Lactobacillus reuteri* probiotics, for paediatric acute diarrhoeal disease in Botswana: A pilot, randomized, factorial, controlled trial. PLoS One. 2017;12(10):e0185177; doi:10.1371/journal.pone.0185177.

25. WHO. The treatment of diarrhoea: a manual for physicians and other senior health workers [online]. World Health Organization; 2005. 4th rev:[Available from: https://apps.who.int/iris/handle/10665/43209 (updated 2005; accessed July 6, 2023).

26. Yousuf EI, Carvalho M, Dizzell SE, et al. Persistence of Suspected Probiotic Organisms in Preterm Infant Gut Microbiota Weeks After Probiotic Supplementation in the NICU. Front Microbiol. 2020;11:574137; doi:10.3389/fmicb.2020.574137.

27. Stearns JC, Davidson CJ, McKeon S, et al. Culture and molecular-based profiles show shifts in bacterial communities of the upper respiratory tract that occur with age. ISME J. 2015;9(5):1246–59; doi:10.1038/ismej.2014.250.

28. Guitor AK, Raphenya AR, Klunk J, et al. Capturing the Resistome: a Targeted Capture Method To Reveal Antibiotic Resistance Determinants in Metagenomes. Antimicrob Agents Chemother. 2019;64(1); doi:10.1128/AAC.01324-19.

29. Guitor AK, Yousuf EI, Raphenya AR, et al. Capturing the antibiotic resistome of preterm infants reveals new benefits of probiotic supplementation. Microbiome. 2022;10(1):136; doi:10.1186/s40168-022-01327-7.

30. Alcock BP, Huynh W, Chalil R, et al. CARD 2023: expanded curation, support for machine learning, and resistome prediction at the Comprehensive Antibiotic Resistance Database. Nucleic Acids Res. 2023;51(D1):D690–d9; doi:10.1093/nar/gkac920.

31. Raphenya AR. RGI [online]. github; 2022. Available from: https://github.com/arpcard/rgi (accessed July 26, 2022).

32. Bar-Zeev N, Moss WJ. Hope and Humility for Azithromycin. The New England journal of medicine. 2019;380(23):2264–5; doi:10.1056/NEJMe1906459.

33. Pop M, Walker AW, Paulson J, et al. Diarrhea in young children from low-income countries leads to large-scale alterations in intestinal microbiota composition. Genome Biology. 2014;15(6):R76; doi:10.1186/gb-2014-15-6-r76.

34. Mizutani T, Aboagye SY, Ishizaka A, et al. Gut microbiota signature of pathogen-dependent dysbiosis in viral gastroenteritis. Sci Rep. 2021;11(1):13945; doi:10.1038/s41598-021-93345-y.

35. David LA, Weil A, Ryan ET, et al. Gut microbial succession follows acute secretory diarrhea in humans. mBio. 2015;6(3):e00381–15; doi:10.1128/mBio.00381-15.

36. Chung The H, Le SH. Dynamic of the human gut microbiome under infectious diarrhea. Curr Opin Microbiol. 2022;66:79–85; doi:10.1016/j.mib.2022.01.006.

37. van Schaik W. The human gut resistome. Philos Trans R Soc Lond B Biol Sci. 2015;370(1670):20140087; doi:10.1098/rstb.2014.0087.

38. Tapela K, Rahube T. Isolation and antibiotic resistance profiles of bacteria from influent, effluent and downstream: A study in Botswana. African Journal of Microbiology Research. 2019;13(15):279–89; doi:10.5897/ajmr2019.9065.

39. Xiang Y, Wu F, Chai Y, et al. A new plasmid carrying *mphA* causes prevalence of azithromycin resistance in enterotoxigenic *Escherichia coli* serogroup O6. BMC Microbiol. 2020;20(1):247; doi:10.1186/s12866-020-01927-z.

40. de Vries LE, Valles Y, Agerso Y, et al. The gut as reservoir of antibiotic resistance: microbial diversity of tetracycline resistance in mother and infant. PLoS One. 2011;6(6):e21644; doi:10.1371/journal.pone.0021644.

41. Anand Paramadhas BD, Tiroyakgosi C, Mpinda-Joseph P, et al. Point prevalence study of antimicrobial use among hospitals across Botswana; findings and implications. Expert Review of Anti-infective Therapy. 2019;17(7):535–46; doi:10.1080/14787210.2019.1629288.

42. Dethlefsen L, Relman DA. Incomplete recovery and individualized responses of the human distal gut microbiota to repeated antibiotic perturbation. Proc Natl Acad Sci U S A. 2011;108 Suppl 1(Suppl 1):4554–61; doi:10.1073/pnas.1000087107.

43. Rogawski McQuade ET, Brennhofer SA, Elwood SE, et al. Frequency of bystander exposure to antibiotics for enteropathogenic bacteria among young children in low-resource settings. Proc Natl Acad Sci U S A. 2022;119(36):e2208972119; doi:10.1073/pnas.2208972119.

44. Luby SP, Rahman M, Arnold BF, et al. Effects of water quality, sanitation, handwashing, and nutritional interventions on diarrhoea and child growth in rural Bangladesh: a cluster randomised controlled trial. Lancet Glob Health. 2018;6(3):e302–e15; doi:10.1016/S2214-109X(17)30490-4.

45. Null C, Stewart CP, Pickering AJ, et al. Effects of water quality, sanitation, handwashing, and nutritional interventions on diarrhoea and child growth in rural Kenya: a cluster-randomised controlled trial. Lancet Glob Health. 2018;6(3):e316–e29; doi:10.1016/S2214-109X(18)30005-6.

46. Grembi JA, Lin A, Karim MA, et al. Effect of water, sanitation, handwashing and nutrition interventions on enteropathogens in children 14 months old: a cluster-randomized controlled trial in rural Bangladesh. J Infect Dis. 2020; doi:10.1093/infdis/jiaa549.

47. Gakidou E, Oza S, Vidal Fuertes C, et al. Improving Child Survival Through Environmental and Nutritional InterventionsThe Importance of Targeting Interventions Toward the Poor. JAMA. 2007;298(16):1876–87; doi:10.1001/jama.298.16.1876.

