## Supplementary material for "Minimal impact on the resistome of children in Botswana after azithromycin treatment for acute severe diarrhoeal disease": Figure S1

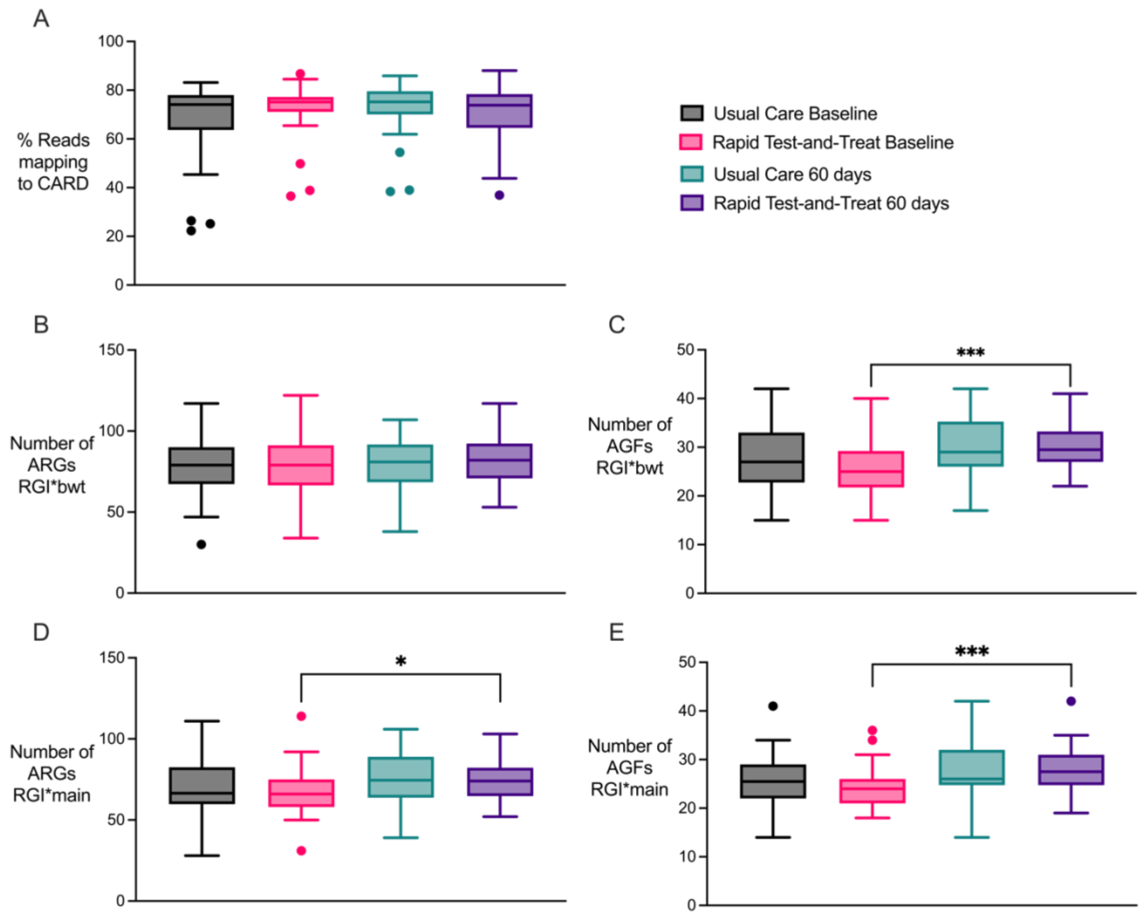


**Supplementary Figure 1: Capturing antibiotic resistance genes in the gut microbiome.** DNA from stool samples collected at baseline and 60 days later was enriched for antibiotic resistance genes prior to sequencing. A) Reads were mapped to CARD and the percentage of reads mapping to CARD in each child was determined. The number of ARGs (B) and the number of AGFs (C) per infant with at least 85% length coverage by at least 50 reads or 100% length coverage by at least 10 reads as determined through RGI*bwt. Paired t-test between RTT Baseline and RTT 60 days: $p < 0.001$. The number of ARGs (D) and the number of AGFs (E) per infant identified as Perfect or Strict hits as determined through *de novo* assembly and RGI*main. Paired t-test between RTT Baseline and RTT 60 days: $p = 0.013$ (D) and $p < 0.001$ (E).

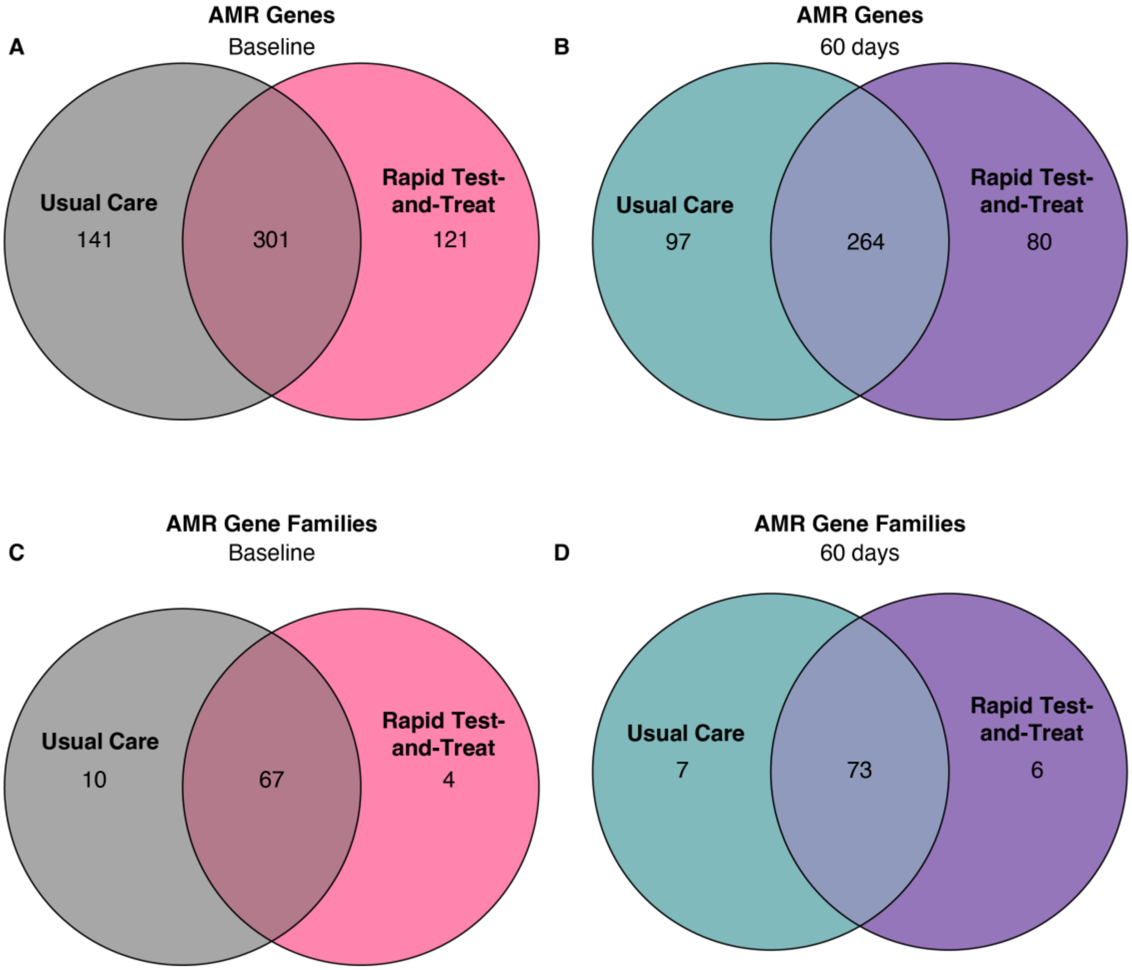


**Supplementary Figure 2:** Unique AMR genes and AMR gene families identified in each cohort at baseline and 60 days post-treatment. This is from the RGI*bwt results.

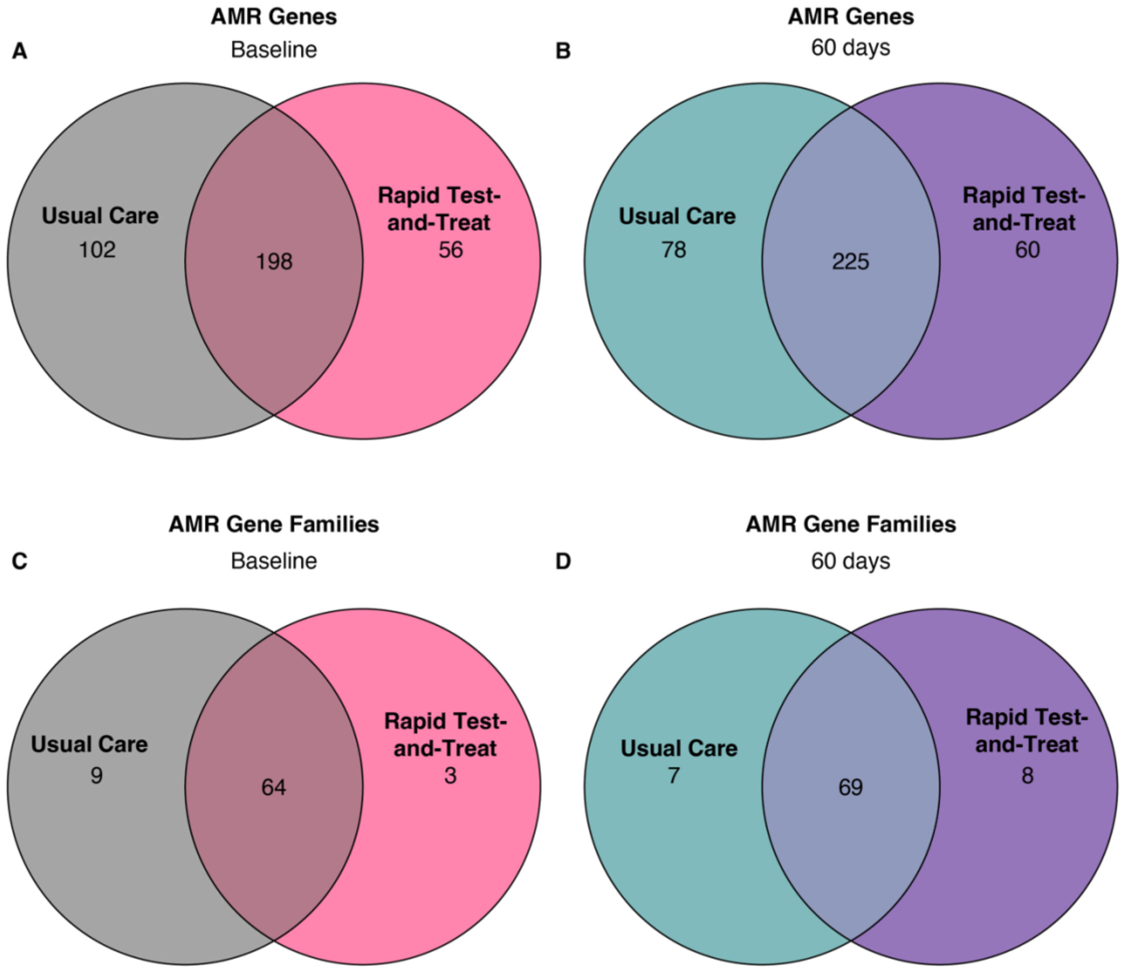


**Supplementary Figure 3:** Unique AMR genes and AMR gene families identified in each cohort at baseline and 60 days post-treatment. This is from the RGI*main results.

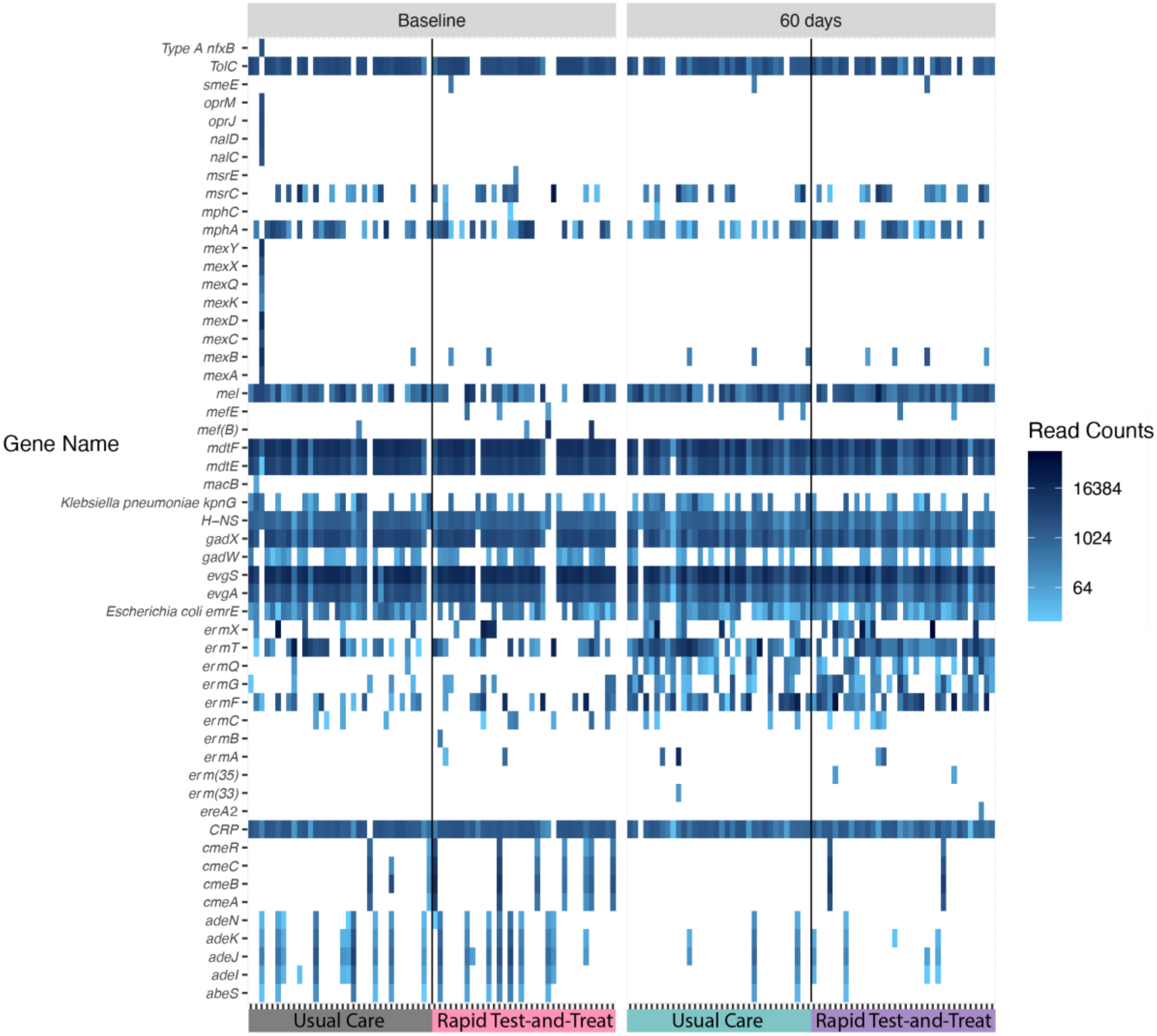


**Supplementary Figure 4**: **Abundance of macrolide resistance genes in infants and changes at 60 days.** Heatmap showing the number of reads mapping to each ARG as determined through RGI*bwt.

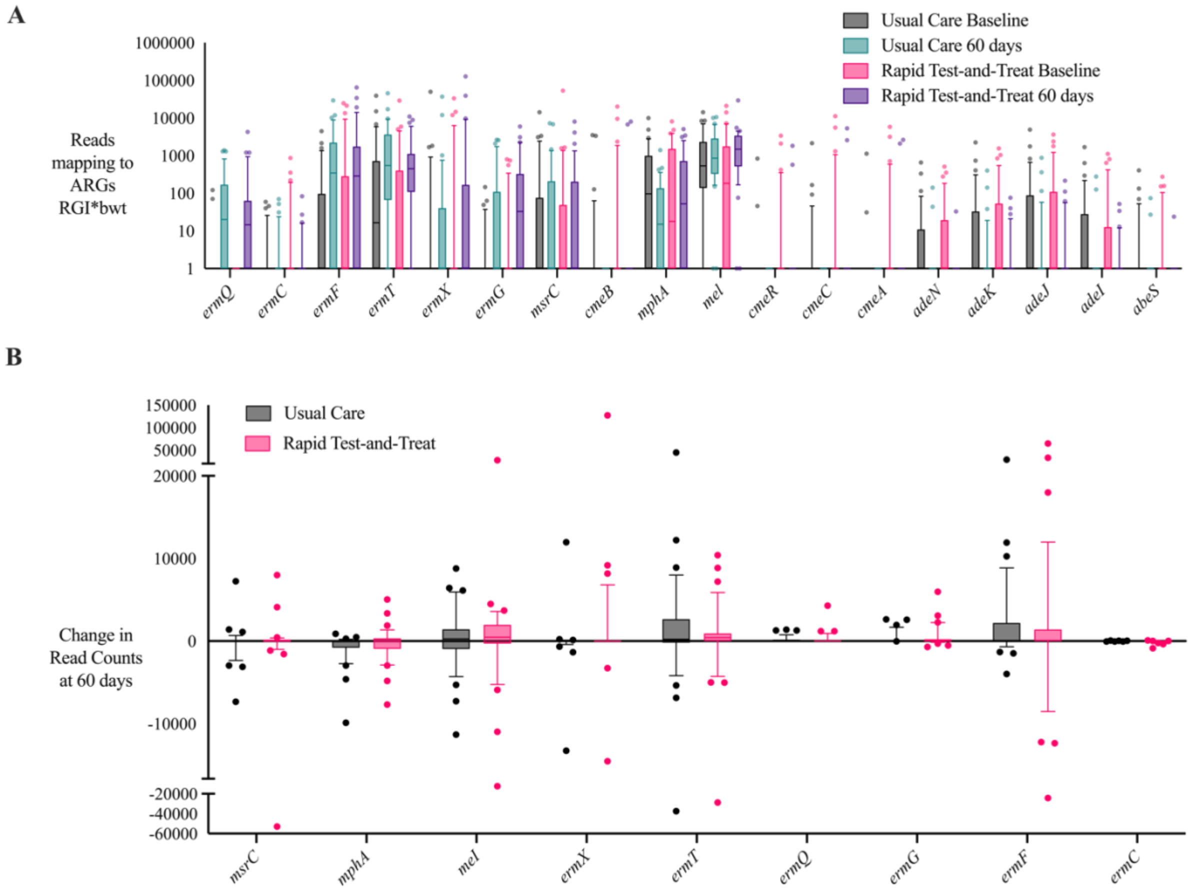


**Supplementary Figure 5: Differences in read count abundance for a subset of ARGs associated with macrolide resistance.** Results were obtained from read mapping to CARD using RGI*bwt. A) Each point represents the number of reads mapping to the specific ARG in one child separated by cohort and timepoint. This set of genes does not include all ARGs associated with macrolide resistance, only the subset of genes that changed in prevalence or abundance in either group of children over time. B) Each point represents the change in read counts mapping to the specific ARG in one child from baseline to 60 days. This plot only encompasses the most prevalent macrolide resistance genes. Box plots represent data within $10^{th}$ to $90^{th}$ percentiles.

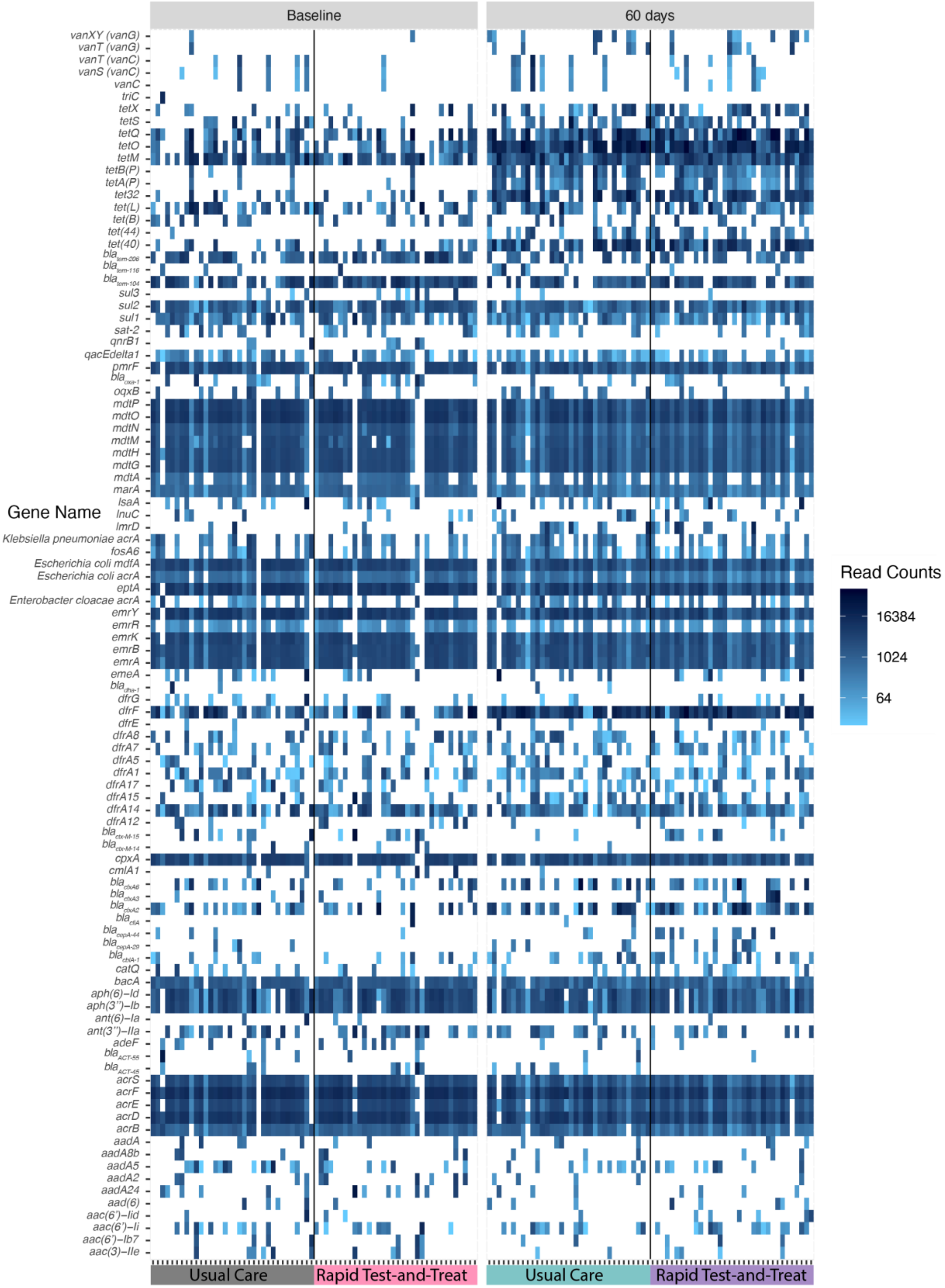

Baseline
60 days
Gene Name
vanXY (vanG)
vanT (vanG)
vanT (vanC)
vanS (vanC)
vanC
triC
tetX
tetS
tetQ
tetO
tetM
tetB(P)
tetA(P)
tet32
tet(L)
tet(B)
tet(44)
tet(40)
bla tem-206
bla tem-116
bla tem-104
sul3
sul2
sul1
sat-2
qnrB1
qacEdelta1
pmrF
bla oxa-1
oqxB
mdtP
mdtO
mdtN
mdtM
mdtH
mdtG
mdtA
marA
lsaA
lnuC
lmrD
Klebsiella pneumoniae acrA
fosA6
Escherichia coli mdfA
Escherichia coli acrA
eptA
Enterobacter cloacae acrA
emrY
emrR
emrK
emrB
emrA
emeA
bla dha-1
dfrG
dfrF
dfrE
dfrA8
dfrA7
dfrA5
dfrA1
dfrA17
dfrA15
dfrA14
dfrA12
bla ctx-M-15
bla ctx-M-14
cpxA
cmlA1
bla cfxA6
bla cfxA3
bla cfxA2
bla cfiA
bla cepA-44
bla cepA-29
bla cblA-1
catQ
bacA
aph(6)–Id
aph(3'')–Ib
ant(6)–Ia
ant(3'')–IIa
adeF
bla ACT-55
bla ACT-45
acrS
acrF
acrE
acrD
acrB
aadA
aadA8b
aadA5
aadA2
aadA24
aad(6)
aac(6')–Iid
aac(6')–Ii
aac(6')–Ib7
aac(3)–IIe
Usual Care
Rapid Test-and-Treat
Usual Care
Rapid Test-and-Treat
Read Counts
16384
1024
64

**Supplementary Figure 6**: **Abundance of top 100 antibiotic resistance genes in infants and changes at 60 days.** Heatmap showing the number of reads mapping to each ARG as determined through RGI*bwt. These were the top 100 ARGs based on overall read abundance across all children and timepoints. ARGs associated with macrolide resistance were omitted and can be found in Supplementary Figure 4. Certain names were shortened; See Additional File 2 for full names and ARO accession numbers.

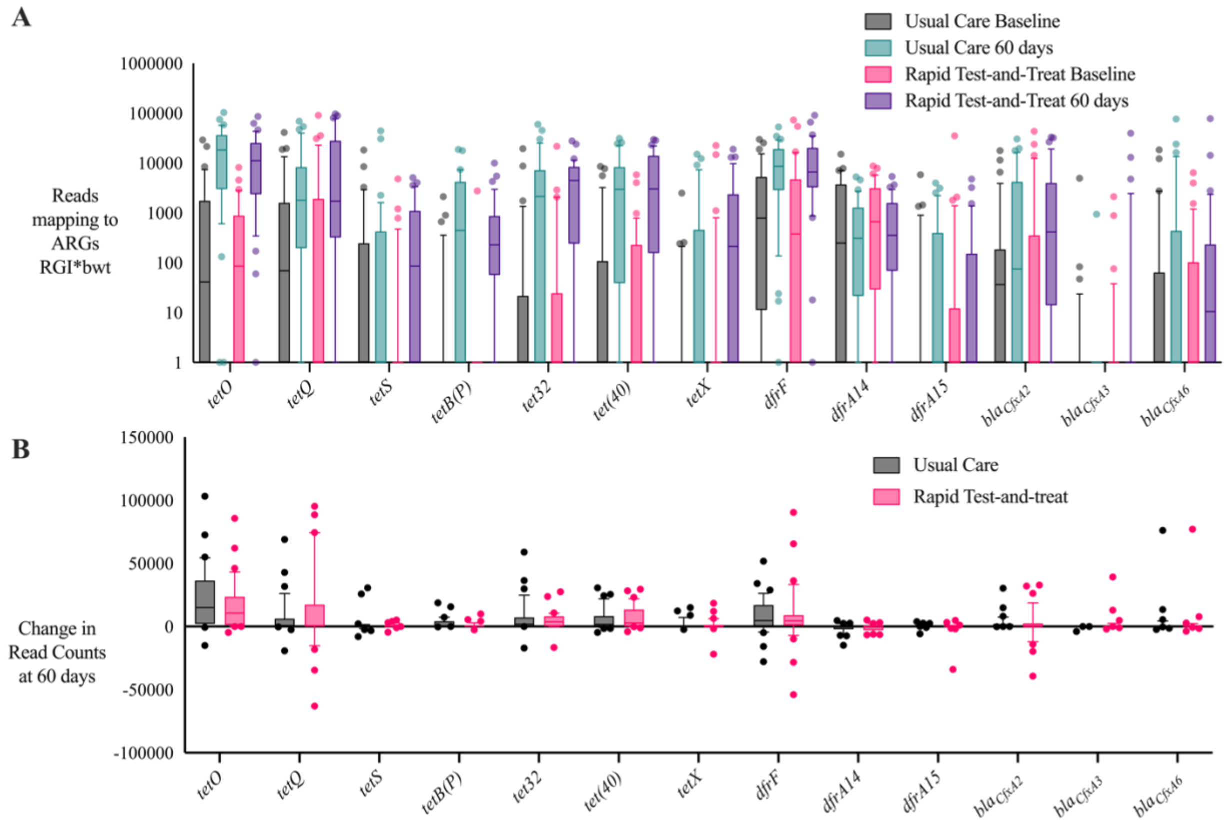


**Supplementary Figure 7: Differences in read count abundance for a subset of ARGs.** Results were obtained from read mapping to CARD using RGI*bwt. A) Each point represents the number of reads mapping to the specific ARG in one child separated by cohort and timepoint. B) Each point represents the change in read counts mapping to the specific ARG in one child from baseline to 60 days. This figure presents a subset of genes that changed in prevalence or abundance in at least one group of children over time. Box plots represent data within $10^{th}$ to $90^{th}$ percentiles.

**Additional File 1:** Participant characteristics and resistome results. Details on study participants including pathogen test results, antibiotic exposure, and sample availability. Table also includes information on DNA library preparation for each sample, sequencing results, and number of ARGs and AMR GFs identified in each sample.

**Additional File 2:** Resistome results from RGI*bwt. Number of reads mapping to each ARG in CARD across all participant samples.

**Additional File 3:** Resistome results from RGI*main. Presence of absence of ARGs as determined through RGI*main analysis for all participant samples. A value of 1 signifies this gene was detected, whereas a value of 0 indicates it was not detected in that participant's sample.

**Additional File 4:** Genetic context of select ARGs. From the RGI*main results, the contigs containing certain ARGs were further analysed for the present of other ARGs, the top blastN hit for the entire contig, and the top taxonomic hit from RGI*kmer_query.

**Additional File 5:** Resistome results from RGI*bwt for negative controls. Number of reads mapping to each ARG in CARD across all sequenced negative controls.

**Additional File 6:** Resistome results from RGI*main for negative controls. Presence of absence of ARGs as determined through RGI*main analysis for all sequence negative controls. A value of 1 signifies this gene was detected, whereas a value of 0 indicates it was not detected in that participant's sample.
