## Supplementary Methods for "Minimal impact on the resistome of children in Botswana after azithromycin treatment for acute severe diarrhoeal disease"

### SUPPLEMENTARY MATERIAL - METHODS

#### DNA extraction and Library preparation

The previously described DNA extraction method involved mechanical lysis with 2.8 mm ceramic beads, enzymatic lysis with lysozyme, proteinase K, and RNase A, followed by phenol-chloroform extraction and purification using the Zymo DNA Clean and Concentrator 25-kit (1, 2). The DNA was quantified using a NanoDrop 2000c Spectrophotometer (Thermo Scientific, Mississauga, ON Canada) and Qubit 1X dsDNA high sensitivity assay, and the quality of the DNA extract was assessed via agarose gel electrophoresis. When available, up to 500 ng of dsDNA was used for input into library preparation with the NEBNext Ultra II dsDNA library kit (Additional File 1). Most libraries received 5 rounds of indexing PCR (Additional File 1). Ten negative controls consisting of a buffer-only extraction blank were included to account for potential contamination from reagents or the laboratory environment. These were processed in the same fashion as the stool DNA extracts, except that 10 rounds of indexing PCR amplification were performed during library preparation due to their low- input.

#### Library enrichment and sequencing

Enrichment for ARGs was performed as previously described (3, 4). To normalize the DNA input for in-solution targeted capture, a High Sensitivity DNA ScreenTape Analysis (Agilent Technologies) was performed to estimate the concentration of each library (Additional File 1). All samples were enriched for 24 hours at 65°C using a previously described probe set to target over 2,000 ARGs (3). Enriched libraries were quantified by quantitative PCR (qPCR), pooled, then sequenced by the Farncombe

Metagenomics sequencing facility at McMaster University on an Illumina MiSeq with 2 x 300 bp sequencing chemistry to a targeted depth of 250,000 clusters per library.

**Analysis of captured antibiotic resistance genes**

Demultiplexed reads were trimmed using skewer v 0.2.2 and string deduplicated using bbtools (dedupe.sh) (5, 6). Reads were subsampled to 150,000 clusters or 300,000 reads using seqtk (v1.3-r117-dirty) (7). Reads were mapped to the Comprehensive Antibiotic Resistance Database (CARD) v 3.2.1 (4891 sequences) using the Resistance Gene Identifier (RGI)*bwt (v 5.2.0) with kma version 1.3.4 (8-10). ARGs reported were filtered for those with at least 85% length coverage of the gene with a least 50 reads mapped or 100% length coverage of the gene with at least 10 reads mapped. Reads were *de novo* assembled using SPAdes v 3.13.0 (metaspades default option), and ARGs predicted using RGI*main v 5.2.0 with CARD v 3.2.1 (using BLAST and with the --exclude_nudge and --low-quality flags included) (11). The potential bacterial hosts of ARGs were predicted using RGI's beta feature for the k-mer prediction of pathogen of origin (RGI*kmer_query) with the default 61-mer database.

We compared results between the two treatment groups at the ARG and the AMR gene family (AGF) levels, which is a higher classification of ARGs. For example, *ermF* (ARO: 3000498) and *ermG* (ARO: 3000522) are both members of the "Erm 23S ribosomal RNA methyltransferase" AGF in CARD. The prevalence of ARGs associated with macrolide resistance, as well as 163 non-macrolide ARGs that were present in at least 10% of children in either treatment group at either timepoint, was determined from the RGI*main results. The $\log_2$ fold change in prevalence of an ARG was calculated using:

$log_2$([prevalence at 60 days]/[prevalence at baseline]). The number of reads mapping to a given ARG was used as an approximation for the abundance of that ARG, given that all samples were subsampled to the same depth (number of reads) and there were no significant differences in the percentage of reads mapping to CARD between groups and time points (Figure S1A). The abundance differences for ARGs associated with macrolide resistance and the top 100 ARGs based on the total number of reads mapping across all samples were determined from RGI*bwt results. The RGI*main and RGI*bwt results almost always reported the same AGFs; however, different variants may be reported at the ARG level, given the nature of both analysis approaches. For example, RGI*main reported TEM-1 and TEM-116, while RGI*bwt predominantly detected TEM-206 and TEM-104. The difference amongst these alleles is 1-5 SNPs over 681 bp, making RGI*bwt susceptible to the allele network problem during read alignment (12).

For the 23S rRNA methyltransferases that confer resistance to macrolides (*ermFGQTX*, AROs: 3000498, 3000522, 3000593, 3000595, 3000596) in which there were appreciable increases in prevalence or differences in abundance between groups and at 60 days, the prevalence of predicted bacterial hosts with the greatest number of k-mer hits from the RGI*kmer_query was compared across cohorts and timepoints. The surrounding genetic context of these methyltransferases, along with the macrolide phosphotransferases (*mphAC,* AROS:3000316, 3000319), was predicted using Prokka version 1.14.5, and the potential origin inferred from the hit with the greatest query coverage and highest percent identity from Nucleotide BLAST (blastN) results against the nonredundant nucleotide collection in NCBI (access Nov 29, 2022) (13-15). Representative contigs showing the diversity of genes surrounding these ARGs were visualized using clinker version 0.0.25 (16).

**Negative controls**

After enrichment, the libraries were analyzed with a High Sensitivity DNA ScreenTape Analysis (Agilent Technologies) and quantified by qPCR. Libraries with sufficient concentration were sequenced on an Illumina MiSeq with 2 x 300 bp chemistry separately from the other stool libraries. Sequencing data was analyzed in the same fashion as the stool samples. From the RGI*bwt read mapping, 46 ARGs were identified, while 122 ARGs were identified through *de novo* assembly and RGI*main (Additional Files 5, 6). The most likely source of these ARGs in the negative controls is cross-contamination during DNA extraction and library preparation. Given the variability in the resistome profiles of the negative controls, a contaminated reagent was not likely. To link the contamination seen in the negative controls with the stool samples, we inspected an alignment of contigs containing *emrB* (ARO: 3000074), a gene involved in an antibiotic efflux system present in most stool samples and all negative controls (results not included). Only one negative control (#3) shared 100% similarity for *emrB* with three other samples, that were processed in the same batch and located in nearby wells on the 96-well plate during library preparation. Given the negligible DNA concentration of the negative extractions as well as the additional rounds of amplification during library preparation, even a small amount of cross-contamination would appear enriched in these controls. The higher levels of endogenous DNA in the stool samples and fewer rounds of indexing amplification are not likely to capture the low level of cross-contamination. Finally, the ARGs of interest that we highlight in this study were of low frequency or not detected in the negative controls; therefore, we do not believe that a small amount of cross-contamination between samples impacted our results.

### REFERENCES


1. Stearns JC, Davidson CJ, McKeon S, et al. Culture and molecular-based profiles show shifts in bacterial communities of the upper respiratory tract that occur with age. ISME J. 2015;9(5):1246-59; doi:10.1038/ismej.2014.250.
2. Yousuf EI, Carvalho M, Dizzell SE, et al. Persistence of Suspected Probiotic Organisms in Preterm Infant Gut Microbiota Weeks After Probiotic Supplementation in the NICU. Front Microbiol. 2020;11:574137; doi:10.3389/fmicb.2020.574137.
3. Guitor AK, Raphenya AR, Klunk J, et al. Capturing the Resistome: a Targeted Capture Method To Reveal Antibiotic Resistance Determinants in Metagenomes. Antimicrob Agents Chemother. 2019;64(1); doi:10.1128/AAC.01324-19.
4. Guitor AK, Yousuf EI, Raphenya AR, et al. Capturing the antibiotic resistome of preterm infants reveals new benefits of probiotic supplementation. Microbiome. 2022;10(1):136; doi:10.1186/s40168-022-01327-7.
5. Jiang H, Lei R, Ding S-W, et al. Skewer: a fast and accurate adapter trimmer for next-generation sequencing paired-end reads. BMC Bioinformatics. 2014;15(1):182; doi:10.1186/1471-2105-15-182.
6. Bushnell B. BBMap - Bushnell B. [online]. github; 2022. Available from: https://github.com/BioInfoTools/BBMap (accessed November 30, 2022).
7. Li H. seqtk [online]. github; 2022. Available from: https://github.com/lh3/seqtk (accessed November 22, 2022).
8. Alcock BP, Huynh W, Chalil R, et al. CARD 2023: expanded curation, support for machine learning, and resistome prediction at the Comprehensive Antibiotic Resistance Database. Nucleic Acids Res. 2023;51(D1):D690-d9; doi:10.1093/nar/gkac920.
9. Clausen P, Aarestrup FM, Lund O. Rapid and precise alignment of raw reads against redundant databases with KMA. BMC Bioinformatics. 2018;19(1):307; doi:10.1186/s12859-018-2336-6.
10. Raphenya AR. RGI [online]. github; 2022. Available from: https://github.com/arpcard/rgi (accessed July 26, 2022).
11. Prjibelski A, Antipov D, Meleshko D, et al. Using SPAdes De Novo Assembler. Current Protocols in Bioinformatics. 2020;70(1):e102; doi:https://doi.org/10.1002/cpbi.102.
12. Lanza VF, Baquero F, Martinez JL, et al. In-depth resistome analysis by targeted metagenomics. Microbiome. 2018;6(1):11; doi:10.1186/s40168-017-0387-y.
13. Seemann T. Prokka: rapid prokaryotic genome annotation. Bioinformatics. 2014;30(14):2068-9; doi:10.1093/bioinformatics/btu153.

14. Altschul SF, Gish W, Miller W, et al. Basic local alignment search tool. J Mol Biol. 1990;215(3):403-10; doi:10.1016/s0022-2836(05)80360-2.
15. NCBI. Nucleotide [online]. 2022. Available from: https://www.ncbi.nlm.gov/nucleotide (accessed November 29, 2022).
16. Gilchrist C. Clinker [online]. github; 2022. Available from: https://github.com/gamcil/clinker (accessed November 22, 2022).
